# Simple calculations of direct impact for the initial assessment of the value of primary HIV prevention interventions

**DOI:** 10.1101/2024.07.13.24310366

**Authors:** Geoff P. Garnett, Josha T. Herbeck, Adam Akullian

## Abstract

**Introduction:** Over the course of the HIV pandemic prevention and treatment interventions have reduced HIV incidence but there is still scope for new prevention tools to further control HIV. Studies of the transmission dynamics and cost effectiveness of HIV prevention tools are often done using detailed complex models but there is a role for simpler earlier analyses.

**Methods:** Equations are defined to calculate the cost effectiveness, budget impact, and epidemiological impact of HIV prevention interventions including equations allowing for multiple interventions and heterogeneity in risk across populations. An efficiency ratio of primary HIV prevention and IV treatment as prevention is defined.

**Results:** As HIV incidence declines the number needed to treat to prevent one HIV infection increases. The cost effectiveness of HIV is driven by incidence, along with efficacy, duration, and costs of the intervention. The budget impact is driven by cost, size of the population and coverage achieved, and impact is determined by the effective coverage of interventions. Heterogeneity in risk could in theory allow for targeting primary HIV prevention but current screening tools do not appear to sufficiently differentiate risk in populations where they have been applied.

**Discussion:** Simple calculations provide a tool to readily assess the cost-effectiveness, impact, and budget impact of HIV prevention interventions and can include heterogeneities in risk of HIV acquisition. These calculations provide rough initial estimates that can be compared with more sophisticated transmission dynamic and health economic models.

**Conclusion:** HIV incidence is declining making primary prevention tools less cost effective. If we require prevention to be more cost effective either we need to target primary prevention tools or they need to be less expensive. Simple equations allow for an exploration of the cost effectiveness of HIV interventions but the sensitivity of results to assumptions needs to be tested by comparison with transmission dynamic models.

## Introduction

The course of HIV epidemics has been dramatically altered by the adoption of safe sex practices, the introduction of antiretroviral treatment, and the introduction of biomedical primary prevention interventions such as voluntary medical male circumcision (VMMC). In many high-income countries and across sub-Saharan Africa HIV incidence has declined as interventions have been scaled. Further declines may be possible with the further introduction of new HIV prevention tools. There is a need to consider what value such new products offer in controlling HIV.

When first introduced there was concern that antiviral treatment of HIV could increase the duration for which a person living with HIV could transmit the virus. However, it was shown that in suppressing viral replication antiviral treatment made HIV untransmissible and that treatment was a prevention tool. One can consider treatment as secondary prevention as it reduces onward transmission once infection has been acquired, and that preventing acquisition in the first place is primary prevention.

In evaluating the potential benefits of new primary HIV prevention interventions attention has been given to detailed assumptions about the product profile, patterns of use, and epidemiological context with mathematical models describing the transmission dynamics of HIV(1). The inclusion of predicted changes in the spread of HIV and the patterns of exposure experienced by cohorts add realism and rigor to such analyses, but also add complexity, as does the detailed consideration of the costs of prevention products and their delivery. The inclusion of such details risks conveying a greater sense of accuracy than is warranted given the uncertainties surrounding both the behaviors of the products, their patterns of use, and the influences on HIV epidemiology.

A systematic review of models exploring the cost-effectiveness of HIV prevention interventions (1) restricted their search to publications from the previous 5 years leaving out a long history of modeling HIV vaccine candidates which later showed no efficacy in trials(2–3)(4). Another systematic review identified 12 papers with cost-effectiveness results for HIV vaccines(5). but left out many modeling papers that explored the potential impact of partially effective HIV without attaching costs(6)(7)(8)(9)(10)(11). Early models show a substantial potential impact from partially effective vaccines. However, the subsequent introduction of other interventions which have reduced the incidence of HIV and AIDS deaths reduces the potential impact of such partially effective vaccines. Models have included both generic vaccine properties(10)(7)(6) and properties for specific vaccines including the gp120 vaccine(9), the RV144 vaccine(12)(13), and the 702 vaccine while they were in phase 3 trials(14)(15–16). The results from these models are greatly influenced by the HIV incidence at the time. The role of HIV incidence is also seen in models of vaginal microbicide gels(17), voluntary medical male (18)(19), oral pre-exposure prophylaxis(20–23), preventive vaginal rings(24–25), and injectable pre-exposure prophylaxis(26–27). Whilst the detail represented in these analyses influences specific results and recommendations, the resulting complex parameter space may obscure important insights into the fundamental drivers of the impact and cost-effectiveness of HIV prevention interventions.

It is possible with simple calculations based on patterns of HIV incidence to approximate the cost effectiveness and impact of new prevention products prior to employing more detailed models. The main drivers of the cost effectiveness of HIV prevention interventions are HIV incidence, efficacy, and the cost of procuring and using prevention tools and the interaction of these can be explored in simple equations.

In the following simple equations for the cost effectiveness, impact and affordability of HIV prevention interventions are defined and their behavior illustrated. The comparative scale of treatment and prevention interventions is compared and considerations for the cost-effectiveness of primary prevention to treatment as prevention are provided. Current patterns of incidence for HIV are explored and applied.

## Methods

### Cost-effectiveness, impact, and affordability calculations

The immediate impact and cost-effectiveness of any primary HIV prevention intervention is a function of the expected HIV incidence in the intervention’s absence. The higher the risk of HIV acquisition the fewer the number of people using the prevention method to prevent an infection:

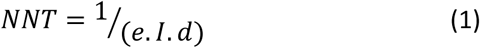

The ‘number needed to treat’ (NNT) is normally calculated comparing the control and intervention arms of trials but can be derived if we know the efficacy of the intervention (*e*), the per susceptible incidence without the intervention (*I*) and the duration of protection.

This number needed to treat (NNT), along with the cost of the intervention determines the cost per HIV infection averted. The cost per infection averted *C* is simply:

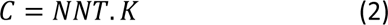

Where *K* is the cost per ‘treatment’. The relevant unit cost of the intervention considered can depend on whose perspective is taken, the program or societies, and whether the average or marginal costs are appropriate.

The above would allow us to compare this cost per infection averted across HIV prevention interventions, but to understand the case for investing in HIV prevention we would need to value the HIV infection averted. With treatment much of the morbidity and mortality associated with HIV infection can be avoided, but the cost of treatment for many years can be compared with the cost of preventing infection in the first place. Offsetting primary prevention cost with treatment costs we can see whether primary prevention can be cost saving. The net costs per infection averted (*net*) are given by:

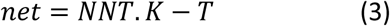

Where T is the cumulative costs of treating an HIV infection.

For comparison with other health interventions not related to HIV we can estimate the disability adjusted life years (DALYs, D) (or alternatively quality adjusted life years (QALYs)) associated with an HIV infection to provide a cost per DALY averted (*Cd*):

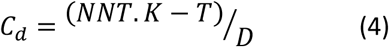

Note that the effective coverage does not influence cost per infection averted but does influence the affordability and the impact of an intervention. Affordability for the provider is determined by the budget impact (*B*) of an intervention. This is the sum costs of scaling up the intervention to a given effective coverage (*f)*:

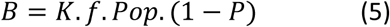

Where *Pop* is the size of the population and *P* is the prevalence of infection (as a proportion). Impact (*A*) the number of new infections averted is the sum of infections averted across the population:

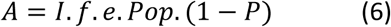

When combining more than one approach to prevention the residual incidence following the effects of one intervention should be applied to the second intervention:

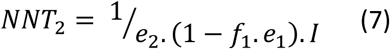

Where *e_j_* and *f_j_* are the efficacy and coverage of intervention *j*. This allows us to calculate the incremental cost effectiveness of the second intervention when added to the first. With multiple interventions then each should be considered in turn with the ‘easiest’ interventions considered first. This assumes that one intervention does not replace another.

In theory heterogeneity in risk allows a different impact and cost effectiveness in sub-populations with different risk of HIV acquisition such that the number needed to treat in risk group *i* (*NNTi*) is:

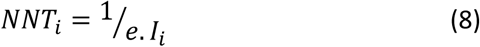

But if coverage includes multiple risk groups, then the relative size of each risk group *fi* and the fraction of those covered in each *gi* matters for the aggregate number needed to treat (NNT) which is a function of the proportion covered falling into each risk group:

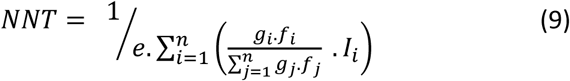

and the coverage of the risk group, *fi*, and actual size of the risk group, Popi, matters for impact:

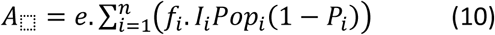

In comparing the cost-effectiveness of treatment as prevention with the cost-effectiveness of primary prevention one can contrast the number of people living with HIV and the number of people susceptible to acquiring HIV. In most populations there will be many more people susceptible than living with HIV. We can see in a simple diagram (Fig. 1) that if 20% of the population are living with HIV and in need of treatment then 80% of the population will be susceptible and be able to benefit from primary prevention.

**Figure 1.**
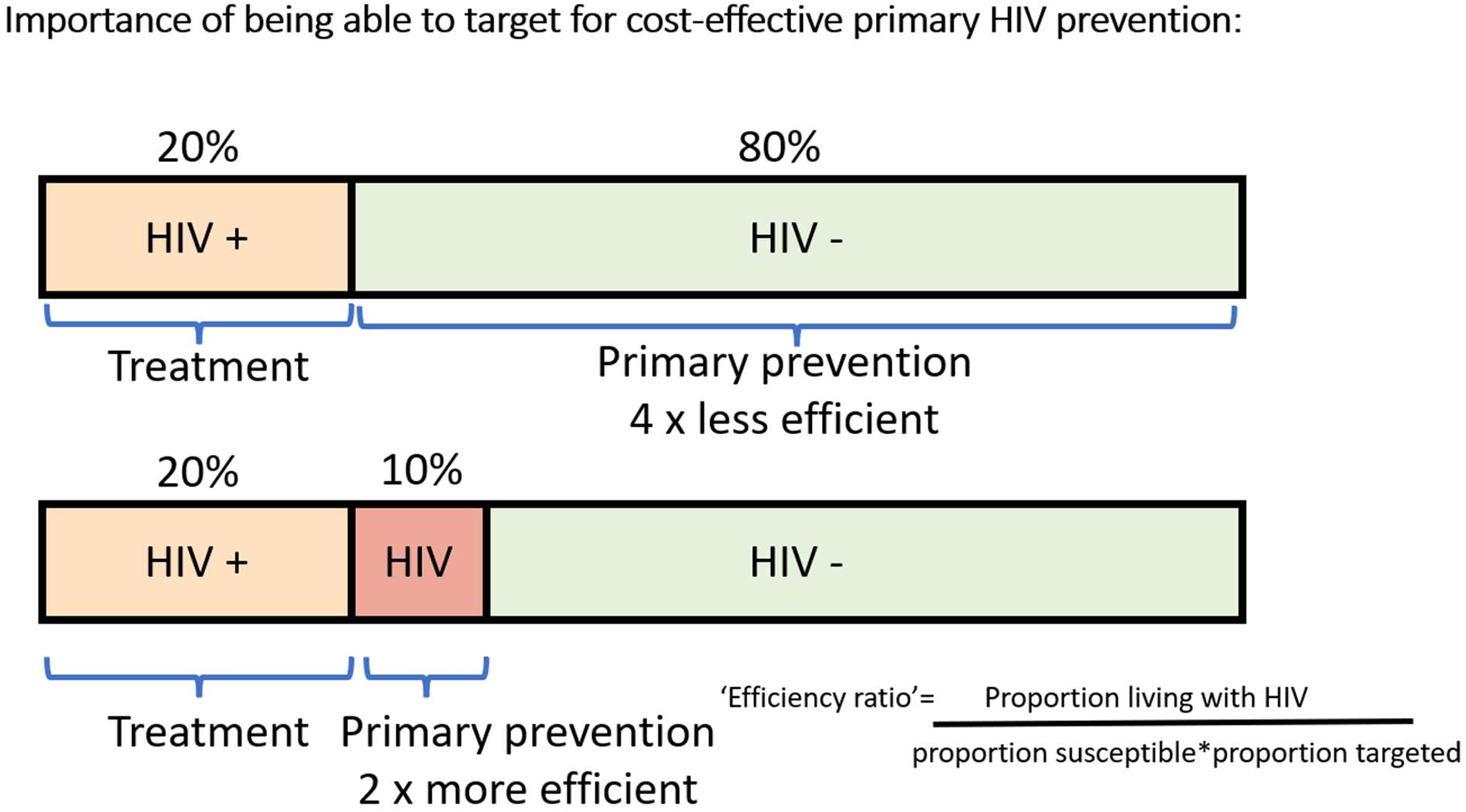
A schematic illustrating the proportion of the population to be covered for treatment as prevention compared with primary HIV prevention.

All other things being equal, for comparable cost effectiveness primary prevention would need to be one quarter of the cost of treatment. If we could target primary prevention to an at-risk fraction, say 10%, of the population then primary prevention could be twice as costly as treatment.

It is possible to think of an efficiency ratio (*E*) between treatment as prevention and primary prevention:

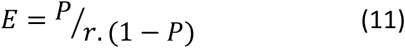

Where r is the fraction of the susceptible population targeted. This efficiency ratio makes assumptions about who living with HIV needs to be treated to avert an HIV infection, and likewise who can benefit from a prevention intervention. Simplistically though, if the number of people living with HIV is greater than the number of susceptible individuals targeted then the cost of primary prevention can be greater than the cost of treatment but remain efficient, whereas when less than one primary prevention needs to cost less. When E equals 1 then treatment as prevention and primary prevention are equivalent.

This does not account for the benefits in reduced morbidity and mortality of treatment for those already living with HIV. The concept of the efficiency ratio could be extended to look at the threshold cost per year of primary prevention (*C_p_*) that would allow it to be more efficient compared to the cost per year of treatment (*C_t_*) and number of years of treatment to avert one DALY (*y*) and the willingness to pay per DALY averted (*W*):

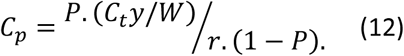

The values derived from these simple equations are explored to generate general insight. A review of estimates of incidence, with publications from 2010 to 2024 identified between 22^nd^ December 2023 and 2^nd^ January 2024, used to generate specific values for numbers needed to treat and the relative efficiency of treatment as prevention and primary prevention. The importance of being able to identify populations that can benefit most from primary prevention is explored in a review of studies looking at risk scores across populations.

## Results

The number needed to treat for a 100% efficacious prevention tool lasting one year is simply the inverse of the per capita HIV incidence in the population using that tool (Eqn. 1). This relationship is shown in Figure 2 along with the same relationship for prevention tools of reduced efficacy.

**Figure 2.**
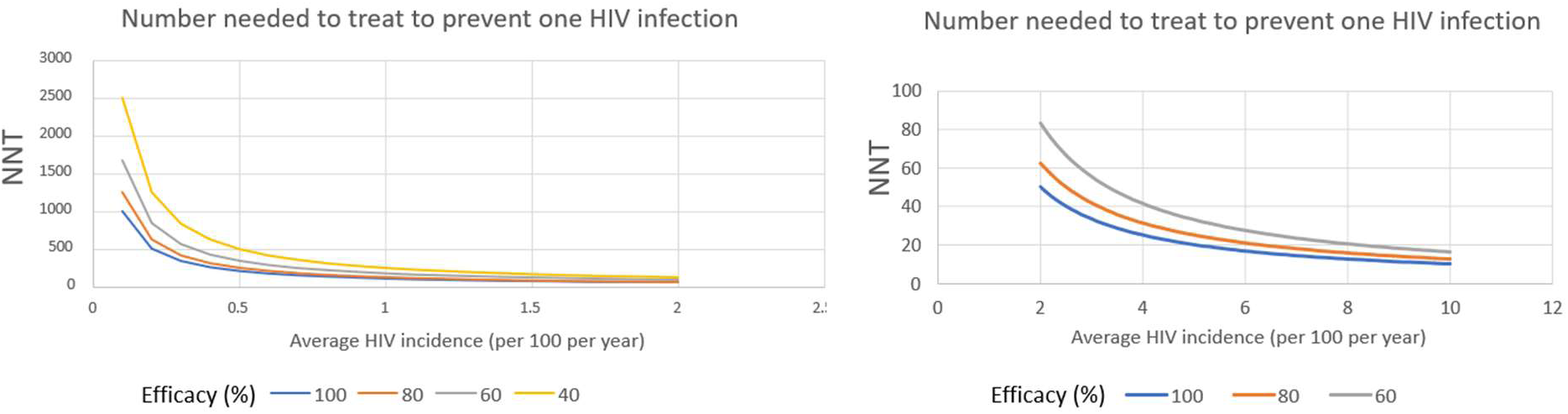
The number needed to treat to prevent one HIV infection as a function of HIV incidence and efficacy of the prevention intervention.

At high incidences of around 5 and 6%, as has been observed in clinical trials then the number needed to treat is in the 10s, but as incidence falls the number rises dramatically into the thousands. To calculate the costs of averting one case of infection one multiplies the number needed to treat by the cost per person using the tool. Thus, for a tool with 100% efficacy costing $1 per user the cost in dollars is simply the number needed to treat, so with a 10% incidence it costs $10 to avert 1 infection, at 1% incidence $100, at 0.1% incidence $1000. These costs per infection averted obviously increase as the cost per prevention tool user increase, so a prevention product such as oral pre-exposure prophylaxis with a cost of $50 per year and an efficacy in users of 50% (efficacy in this case is the product of the biological efficacy and fraction of exposures where there is adherence) in a population with an incidence of 5% would cost $2,000 per infection averted, but in a population with 0.5% incidence would cost $20,000 per infection averted. For the same two incidence rates an injectable pre-exposure prophylactic tool costing $200 per person per year with a 95% efficacy would cost $4,210 and $42,105 per HIV infection averted.

Measuring the costs of HIV treatment has been done in detail in different contexts for example models of treatment in South Africa (28) and across clinics in Uganda and Tanzania (29). The costs of treatment vary greatly across countries and depend on the costs that are included, and for a lifetime of treatment the discount rate assumed. For ease of illustration, we will assume that treatment costs $500 per person per year and is needed for 30 years, so adding to $15,000 for a lifetime. This can be compared with the costs of preventing one infection, favoring prevention if the cost per infection averted is $2,000 but not $20,000. If the costs of treatment exceed the costs of primary prevention, then prevention is cost saving and, in theory dominates. If we have an estimate of the DALYs associated with an HIV infection, say 5, then we can calculate the costs per DALY gained (Eqn. 4), such that for preventing treatment costs of $15,000 with prevention costs of $20,000 we spend $1,000 per DALY, which is slightly more than the willingness to pay per DALY of between $547 and $872 revealed HIV by spending in South Africa (30).

Calculations of Affordability require more information including the size of the at-risk population, coverage, and HIV prevalence. For a population of 1 million with an existing prevalence of 20%, a coverage of susceptible individuals of 50%, a $50 prevention intervention would cost $20,000,000, one of $200 would cost $80,000,000 (Eqn. 5). With a per susceptible incidence of 5% and an efficacy of 50% this would avert 10,000 HIV infections (Eqn. 6)

These straightforward calculations depend on knowing HIV incidence.

### Measuring and estimating incidence

There are essentially 3 ways to measure incidence: 1) through complete enumeration of new HIV diagnoses; 2) through sampling a population and either prospectively measuring new infections in a cohort or retrospectively detecting new infections amongst those living with HIV using a recency assay; 3) or through trends in HIV prevalence and mortality to estimate trends in incidence. All three suffer from biases and errors leading to uncertainty in measures of incidence and therefore uncertainty in the cost-effectiveness of HIV prevention.

Measurement of HIV incidence in population cohorts provides a direct measure of incidence, but most published measures are for specific populations and are a few years out of date. In establishing HIV prevention trial sites in Uganda, Kenya, Rwanda, South Africa, and Zambia incidences measured up to 2013 ranged between 1.4 per hundred person years to 10.8(31). A systematic review of studies published between 2010 and 2019 identified 291 studies covering 22 African countries(32). The analysis found significant declines in HIV incidence over the period. For the general population incidence estimates ranged from 0.35 to 3.28 per hundred person years. Amongst the 5 cohorts studying men who have sex with men (MSM) incidence ranged from 1.03 to 15.4, for the 7 studies of female sex workers it ranged from 0.6 to 3.4. Population based cohorts from Karonga in Malawi, Kisesa in Tanzania, Manicaland in Zimbabwe, Masaka in Uganda, Rakai in Uganda, and uMkhanyakude in South Africa were analyzed as part of the ALPHA network of cohorts and show incidence amongst different age groups of men and women declining over time between 2000 and 2017. In general incidence per 100 person years fell from around a few to less than one. The highest incidence was observed in uMkhanyakude which fell steeply, but late on in the time period of the analysis(33). In evaluating the DREAMS project HIV incidence amongst 15 to 24 year old women in uMkhanyakude in KwaZulu Natal was observed over the period 2016 to 2018 and found to be 2.8 per hundred person years amongst 15-19 year olds and 5.8 amongst 20 to 24 year olds. This compares to the rates observed for Gem in Kenya where the incidence amongst 20 to 24 old women was 0.64 per hundred person years (34). Amongst ECHO trial participants randomized to different contraceptive options in South Africa between 2015 and 2018 incidence was 4.51 (4.05-5.02) per hundred person years(31). Other high-risk populations include men who have sex with men (MSM) in South Africa with an incidence of 6.2% (which was as high as 21.8% in 18-19 year old MSM(31,35))

Exploring the implications of these incidence patterns we can calculate the number needed to cover with HIV prevention to avert one HIV infection. Assuming 100% efficacy 22 women from the ECHO trial in South Africa would need prevention to avert one case of infection, amongst 20-24 year old women in uMkhanyakude it would be 17, whereas for the same age group in GEMS it would be 167. The ranges from the systematic review for the general population would be as high as 285 and as low as 30, for MSM as high as 97 and as low as 6.5, and for female sex workers as high as 166 and as low as 29.

UNAIDS estimates incidence through fitting models to HIV prevalence data from sentinel populations such as pregnant women and population-based surveys(36). Estimates of incidence across all ages for each country have been published by UNAIDS(37). Assuming this incidence occurs amongst adults we can calculate the incidence for those adults by dividing the all-age incidence by the fraction of the population that is adult (from the World Bank estimates). Further we can calculate the incidence amongst those susceptible through dividing by the proportion susceptible (subtracting the prevalence of HIV). This allows us to calculate the number of susceptible individuals needing prevention to avert one HIV infection from each country using equation 1 assuming levels of efficacy (assumed 90% here, and the costs of preventing an HIV infection using equation 2 assuming costs of per person using the prevention intervention, in this case $50 (Table 1).

**Table 1.**
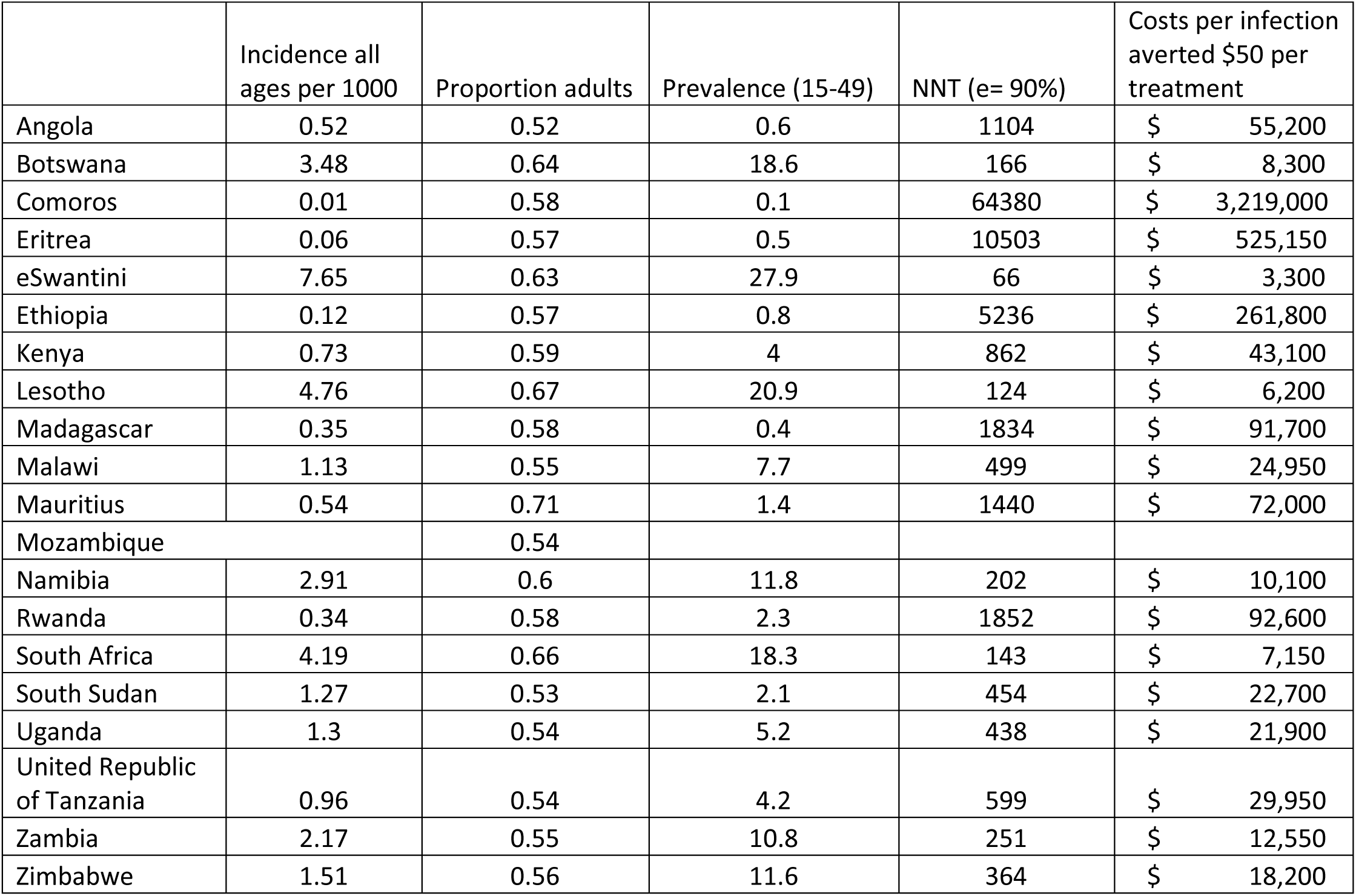
Calculations of the number needed to treat and cost-effectiveness of HIV prevention for African populations based on UNAIDS estimates of HIV incidence.

### Heterogeneity in risk

These studies provide an average incidence over the whole populations. For most general populations now, incidence is below 1% and often below 0.5%. For some specific age groups of women in high-risk locations such as KwaZulu Natal, or amongst young sex workers and MSM it can be much higher. If we can identify who is most at risk in a population then we can focus HIV prevention. However, there is a tradeoff between efficiency and impact. Focusing on everyone is the least cost-effective but most impactful approach. This can be illustrated with a hypothetical population described in Table 2 where there are 4 categories representing different risks of HIV, e.g. 2% of the population with a 5% incidence and 70% with a much lower incidence of 0.01. One can see how the number needed to treat is much lower in the high-risk. In addition, over a longer time frame those with a higher risk could generate most future infections and more infections still can be prevented through prevention in this group. Hence the need for transmission dynamic models.

**Table 2.** Calculations of numbers needed to treat to prevent one HIV infection in a hypothetical heterogeneous population.

| HIV Incidence | NNT | Proportion of the population | Infections per 100,000 |
| --- | --- | --- | --- |
| 5% | 20 | 2% | 100 |
| 1% | 100 | 8% | 80 |
| 0.02% | 5000 | 20% | 4 |
| 0.01% | 10000 | 70% | 7 |

In this hypothetical example we can explore how the distribution of a prevention intervention across the population could influence both the impact and cost-effectiveness of interventions applying equations 9 and 10. For example if in a population of 100,000 prevention with 100% efficacy was used by 1,000 people then if all 1,000 were in the highest risk group then 50 infections would be prevented with an number needed to treat of 20. If the 1,000 was equally distributed across the two highest risk groups then the impact would be 30 infections prevented with an NNT of 33, and if equally distributed across the 3 highest groups 20 infections would be prevented with a NNT of 50.

This though, is a hypothetical example. How well can we describe risk empirically. Some studies have tried to identify risk factors for HIV acquisition and have been able to divide populations according to risk scores depending on these variables(38)(39)(40). Such analyses allow us to estimate the fraction of the population amongst which most infections fall. In the study of Kagaayi and colleagues(38) from Uganda in 2003 to 2011 where incidence rates were 0.98 (0.86-1.12) for men, and 1.11 (1.0-1.24) for women, amongst men 55.1 %, 23.6% and 22.3% of infections and for women 48.0%, 20.7% and 31.3% of infections were in the top quarter, the second quarter and bottom half of the population according to risk scores This would give average NNT for men of 102 and for women of 90 but NNTs can be calculated for different parts of the population (Table 3).

**Table 3.** Observed heterogeneity in HIV incidence in a Ugandan population and the associated number needed to treat to prevent one HIV infection.

| Fraction of infections men | HIV incidence (%) men | NNT men | Fraction of infections women | HIV incidence (%) women | NNT women |
| --- | --- | --- | --- | --- | --- |
| 0.551 | 2.16 | 46 | 0.48 | 2.13 | 47 |
| 0.236 | 0.93 | 108 | 0.267 | 0.92 | 109 |
| 0.213 | 0.42 | 240 | 0.313 | 0.69 | 144 |

Looking at the data presented by Balkus et al(39) for the VOICE study population of women between 2009 and 2011 we can see the proportion of HIV infections falling in a fraction of the population defined by risk scores (Fig. 3).

**Figure 3.**
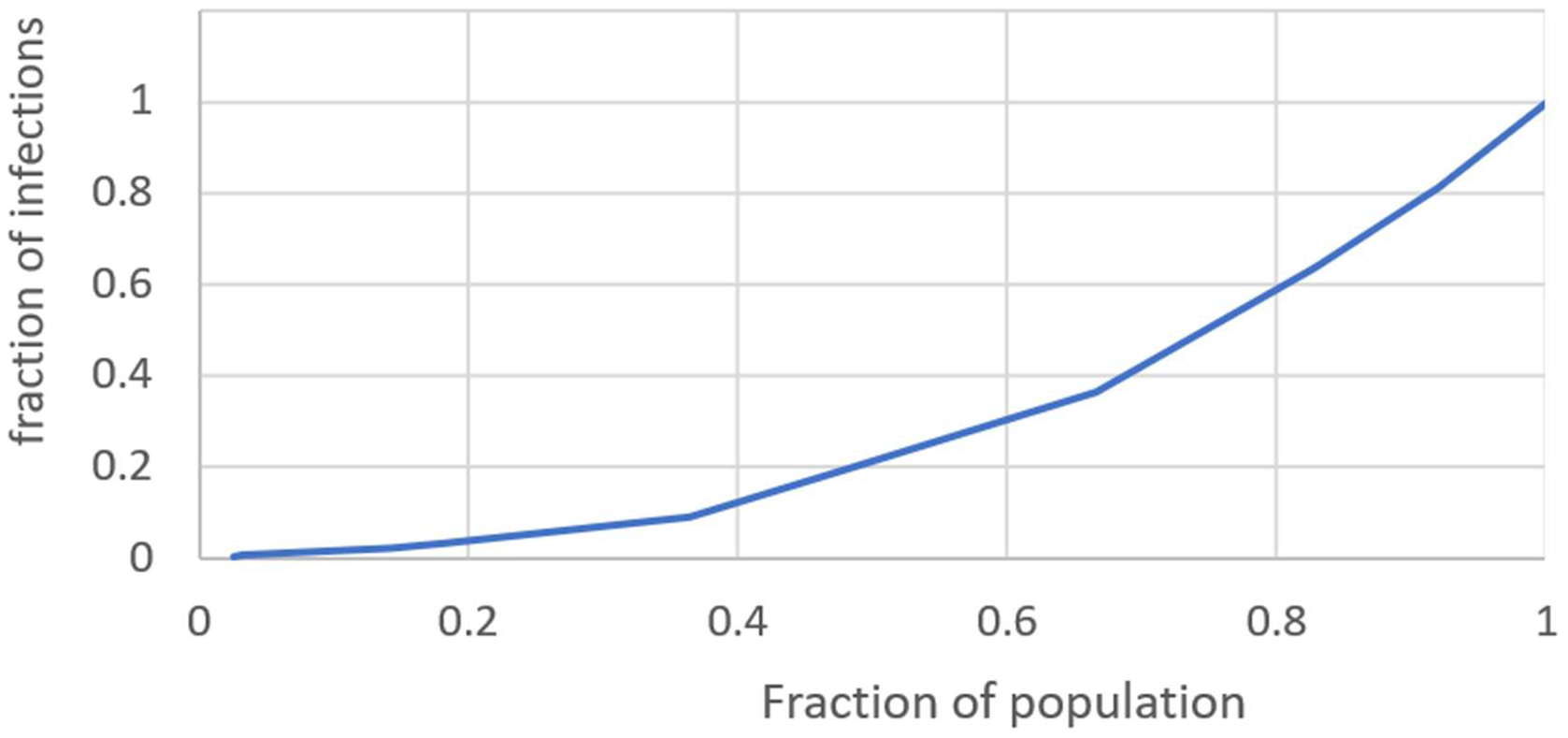
The fraction of infections falling within a given fraction of the VOICE trial population.

The number needed to treat to prevent an HIV infection and the proportion of infections falling within each category can be calculated (Table 4)

**Table 4.**
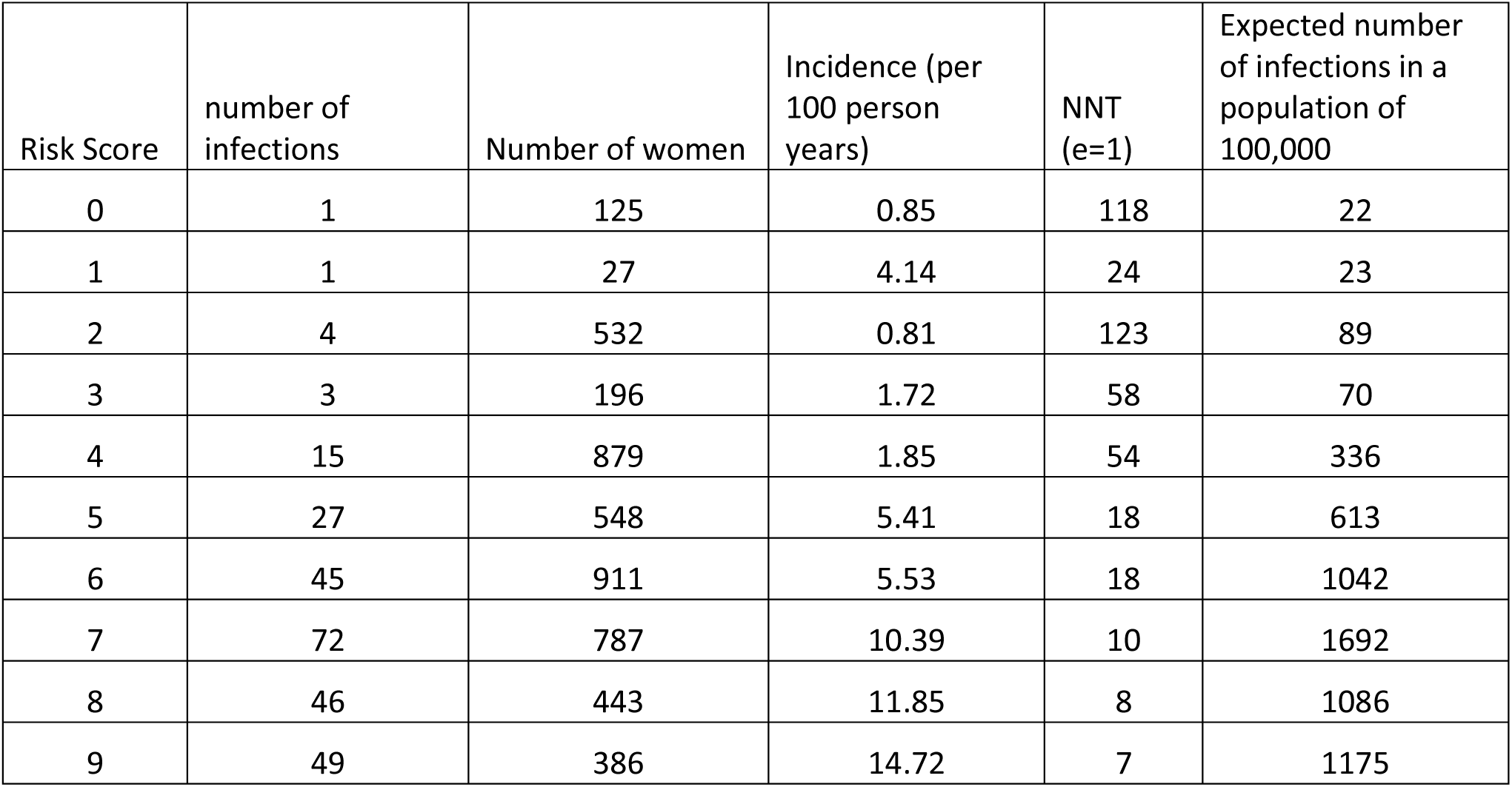
Observed heterogeneity in HIV incidence observed in the Voice trial and associated numbers needed to treat to prevent one HIV infection.

| Risk Score | number of infections | Number of women | Incidence (per 100 person years) | NNT (e=1) | Expected number of infections in a population of 100,000 |
| --- | --- | --- | --- | --- | --- |
| 0 | 1 | 125 | 0.85 | 118 | 22 |
| 1 | 1 | 27 | 4.14 | 24 | 23 |
| 2 | 4 | 532 | 0.81 | 123 | 89 |
| 3 | 3 | 196 | 1.72 | 58 | 70 |
| 4 | 15 | 879 | 1.85 | 54 | 336 |
| 5 | 27 | 548 | 5.41 | 18 | 613 |
| 6 | 45 | 911 | 5.53 | 18 | 1042 |
| 7 | 72 | 787 | 10.39 | 10 | 1692 |
| 8 | 46 | 443 | 11.85 | 8 | 1086 |
| 9 | 49 | 386 | 14.72 | 7 | 1175 |

Being able to focus on those most at risk is necessary for primary prevention to be more efficient than treatment as prevention. Figure 4 illustrates the Efficiency ratio for a range HIV prevalence and fraction of the susceptible population to be targeted.

**Figure 4.**
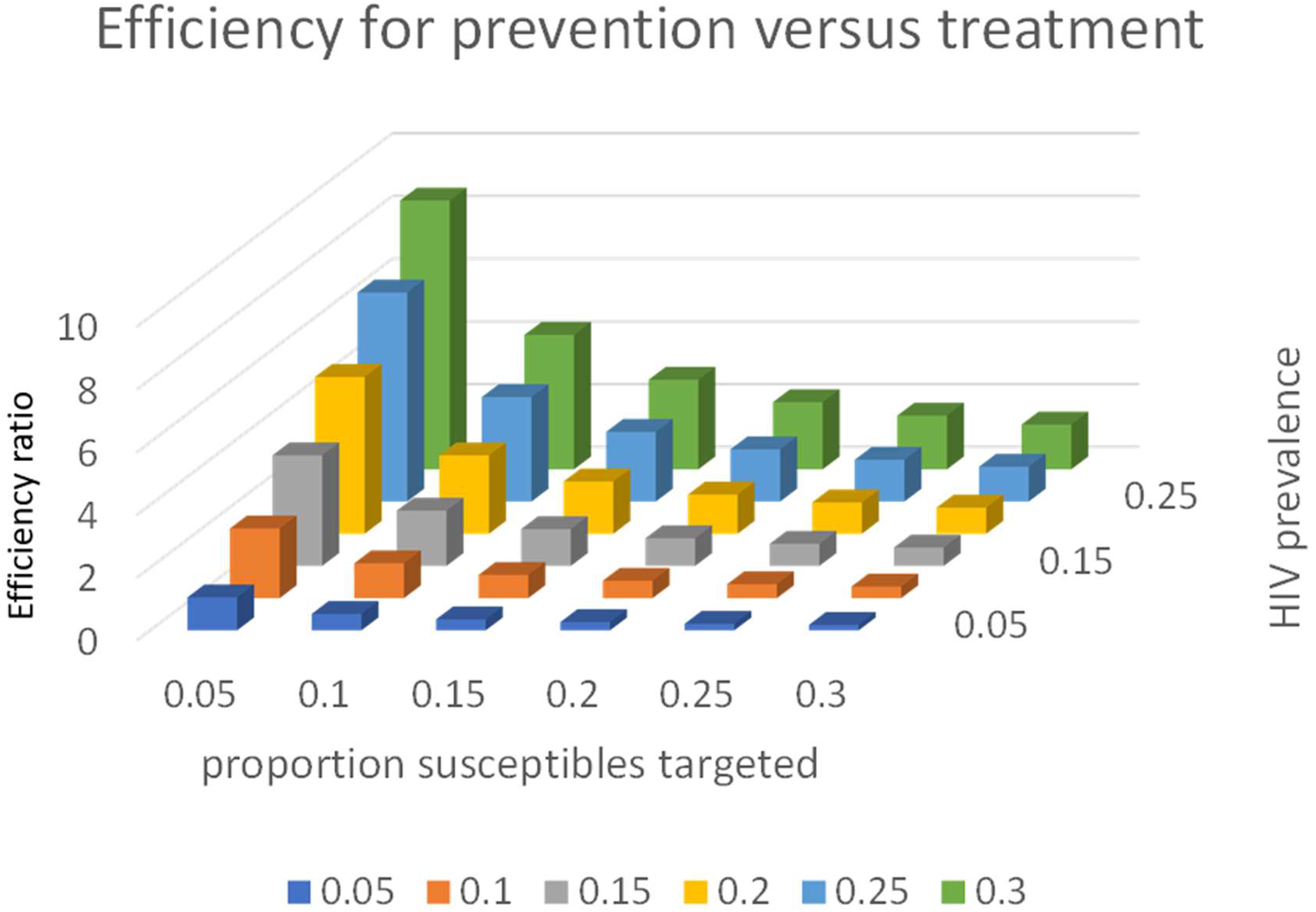
The Effcicacy ratio (eqn 12) for treatment as prevention and primary HIV prevention.

We can calculate the threshold price per person for prevention in Eqn. 12. Assuming 30 years of treatment saves 20 DALYs (the expected 10-year survival expected without the treatment and 20 added years life expectancy through treatment), a cost per year of treatment of $500 and a willingness to pay per DALY of $750) then with an HIV prevalence of 20% if all susceptible individuals need to receive prevention then the cost for prevention can only be $0.25 per year. If only 1% of susceptible individuals need prevention this increases to $25. To increase this threshold the costs of treatment or the willingness to pay to gain a DALY need to increase. The cost of treating those currently missing from care may indeed be much greater than the current costs of treatment.

## Discussion

We have described simple equations for calculating costs effectiveness, budget impact and epidemiological impact for HIV prevention interventions. The illustration of these calculations shows the importance of incidence for cost effectiveness. With current estimates of HIV incidence across population the number needed to treat to prevent one HIV infection is becoming large and to be cost-effective either prevention tools will need a low price or to be targeted at those with a relatively high risk.

Heterogeneity in HIV incidence should allow for targeting HIV interventions but there are questions about how feasible targeting is. Are there variables or circumstances we can use to identify those who can benefit more from HIV prevention tools? Before programming HIV prevention can we measure patterns of HIV incidence or would this be prohibitively expensive. Research is needed to identify generalizable risk factors beyond standard geographic and demographic variables. If these can be identified then we can cost effectively target primary HIV prevention, if they prove elusive then we will struggle to find a substantial role for expensive primary prevention.

There is a question about the precision and accuracy needed to help policy makers make better decisions. When is confidence in results sufficient to allow decisions and what is the value of providing more precise results? The more expensive HIV prevention tools are, the more accurately they will need to be targeted to be cost effective. However, there is a cost to generating the information needed to target HIV prevention interventions, and we should consider when this cost outweighs the improved efficiency that is possible.

Our analyses makes simplifying assumptions in calculating epidemiological impacts and calculating costs. One of the most important is the lack of indirect effects of the interventions. Preventing the acquisition of an infection averts not only the infection and disease in the person directly benefiting but also other subsequent infections that would derive from that infection. This means that calculations of benefits amongst those directly protected underestimate the effects of prevention. It is most important to consider the influence of transmission dynamics on cost effectiveness when the qualitative conclusions from calculation could be altered by the lack of their inclusion. Because HIV epidemics play out over decades rather than weeks and months the nonlinear dynamics of HIV have less influence on health economic analyses than those for many short-lived infections diseases, especially if short time horizons are used to consider costs and benefits. If the cost effectiveness of HIV prevention is clear from our analysis then it is unlikely that transmission dyanamics will change the qualitative conclusions, but as decisions become more marginal then the more important a role they could play. More work is needed to systematically compare linear models of HIV control with transmission dynamics models to better understand when the former are adequate.

In terms of HIV prevention costs. If the intervention is delivered through an existing system, then the marginal costs of adding the prevention intervention are likely appropriate. Whereas, if the entire system to deliver the intervention must be built from scratch, then the average cost is more appropriate.

Over the course of the HIV pandemic incidence of HIV infection has variously been reduced by the saturation of infection in sub-populations, reductions in numbers of sexual partners and condom use, voluntary medical male circumcision, and treatment as prevention. What the right balance is between treatment and primary prevention is, and how to maximize reductions in HIV and AIDS is still a relevant question. The harder it is to find and maintain on treatment those who are still virally unsuppressed the more role there will be for primary prevention. The more inexpensive and easy to use prevention tools are, the more call there will be for their widespread use.

## Conclusions

The main drivers of the cost effectiveness of HIV prevention interventions are HIV incidence, the efficacy and duration of protection from a prevention product, and the cost of the use of that product. Simple calculations allow a rapid assessment of the relationship between the characteristics of an HIV prevention product and its cost-effectiveness, impact, and affordability. Use in those with a greater risk makes products more cost effective but our current approaches in sub-Saharan African populations do not allow for substantial gains in efficiency.

## Data Availability

All data produced in the present work are contained in the manuscript

## Authors’ contributions

GPG conceived the paper, developed the equations, performed the calculations, and drafted the manuscript. JH and AA provided input on the equations and calculations and revised the manuscript.

## Acknowledgements

The authors thank the many modelers and health economists they have interacted with over the years exploring the health economics of HIV interventions.

## Disclaimer

The views expressed in this article are those of the authors and do not necessarily represent those of the Bill and Melinda Gates Foundation.

## Data Availability Statement

All the data analyzed in this paper was taken from the published literature.

## References

1. Bozzani FM, Terris-Prestholt F, Quaife M, Gafos M, Indravudh PP, Giddings R, et al. Costs and Cost-Effectiveness of Biomedical, Non-Surgical HIV Prevention Interventions: A Systematic Literature Review. PharmacoEconomics. 2023;41(5):467–80.

2. McLean AR & Blower SM Imperfect vaccines and herd immunity to HIV. Vol. 253, Proceedings of the Royal Society of London. Series B: Biological Sciences. 1993; 253 (136):9–13.

3. Stover J, Bollinger L, Hecht R, Williams C, Roca E. The Impact of An AIDS Vaccine In Developing Countries: A New Model And Initial Results. Vol. 26, Health Affairs. 2007; 26 (4): 1147–58.

4. Harmon TM, Fisher KA, McGlynn MG, Stover J, Warren MJ, Teng Y, et al. Exploring the Potential Health Impact and Cost-Effectiveness of AIDS Vaccine within a Comprehensive HIV/AIDS Response in Low- and Middle-Income Countries. PLOS ONE 2006; 11 (1). e0146387.

5. Adamson B, Carlson J, Kublin J, Garrison L. The Potential Cost-Effectiveness of Pre-Exposure Prophylaxis Combined with HIV Vaccines in the United States. Vaccines 2017; 5 (2): 13.

6. Blower S. Modeling the Potential Public Health Impact of Imperfect HIV Vaccines. Vol. 192, The Journal of Infectious Diseases. 2005; 192 (8): 1494–5.

7. Anderson R, Hanson M. Potential Public Health Impact of Imperfect HIV Type 1 Vaccines. Vol. 191, The Journal of Infectious Diseases 2005; 191 (Suppl. 1): S85–96.

8. Anderson R, Garnett G. Low-efficacy HIV vaccines: potential for community-based intervention programmes. The Lancet 1996; 348 (9033) 1010–3.

9. Anderson RM, Swinton J, & Garnett GP. Potential impact of low efficacy HIV-1 vaccines in populations with high rates of infection. Proceedings of the Royal Society of London. Series B: Biological Sciences. 0. 1995; 261 (136): 147–51.

10. Abu-Raddad LJ, Boily M-C, Self S, Longini IM. Analytic Insights Into the Population Level Impact of Imperfect Prophylactic HIV Vaccines. Journal of Acquired Immune Deficiency Syndromes 2007; 45 (4): 454–67.

11. Rida W, Sandberg S. Modeling the Population Level Effects of an HIV-1 Vaccine in an Era of Highly Active Antiretroviral Therapy. Bulletin of Mathematical Biology 2009: 71 (3): 648–80.

12. Andersson KM, Paltiel AD, Owens DK. The potential impact of an HIV vaccine with rapidly waning protection on the epidemic in Southern Africa: Examining the RV144 trial results. Vaccine 2011; 29 (36) 6107–12.

13. Hontelez JAC, Nagelkerke N, Bärnighausen T, Bakker R, Tanser F, Newell M-L, et al. The potential impact of RV144-like vaccines in rural South Africa: A study using the STDSIM microsimulation model. Vaccine. 2011; 29 (36): 6100–6.

14. de Montigny S, Adamson BJS, Mâsse BR, Garrison LP, Kublin JG, Gilbert PB, et al. Projected effectiveness and added value of HIV vaccination campaigns in South Africa: A modeling study. Scientific Reports. 2018; 8(1): 6066.

15. Selinger C, Dimitrov DT, Welkhoff PA, Bershteyn A. The future of a partially effective HIV vaccine: assessing limitations at the population level. International Journal of Public Health. 2019; 64 (6): 957: 957–64.

16. Selinger C, Bershteyn A, Dimitrov DT, Adamson BJS, Revill P, Hallett TB, et al. Targeting and vaccine durability are key for population-level impact and cost-effectiveness of a pox-protein HIV vaccine regimen in South Africa. Vaccine. 2019; 37 (16):2258–67.

17. Terris-Prestholt F, Foss AM, Cox AP, Heise L, Meyer-Rath G, Delany-Moretlwe S, et al. Cost-effectiveness of tenofovir gel in urban South Africa: model projections of HIV impact and threshold product prices. BMC Infect Dis. 2014;14(1).

18. Kripke K, Reed J, Hankins C, Smiley G, Laube C, Njeuhmeli E. Impact and Cost of Scaling Up Voluntary Medical Male Circumcision for HIV Prevention in the Context of the New 90-90-90 HIV Treatment Targets. PLoS One. 2016;11(10):e0155734.

19. Bansi-Matharu L, Mudimu E, Martin-Hughes R, Hamilton M, Johnson L, Brink Dt, et al. Cost-effectiveness of voluntary medical male circumcision for HIV prevention across sub-Saharan Africa: results from five independent models. Lancet Glob Health. 2023;11(2):e244–55.

20. Case KK, Gomez GB, Hallett TB. The impact, cost and cost-effectiveness of oral pre-exposure prophylaxis in sub-Saharan Africa: a scoping review of modelling contributions and way forward. J Int AIDS Soc. 2019;22(9):.

21. Pretorius C, Schnure M, Dent J, Glaubius R, Mahiane G, Hamilton M, et al. Modelling impact and cost-effectiveness of oral pre-exposure prophylaxis in 13 low-resource countries. J Int AIDS Soc. 2020;23(2):.

22. Gomez GB, Borquez A, Case KK, Wheelock A, Vassall A, Hankins C. The Cost and Impact of Scaling Up Pre-exposure Prophylaxis for HIV Prevention: A Systematic Review of Cost-Effectiveness Modelling Studies. PLoS Med. 2013;10(3):e1001401.

23. Stone J, Bothma R, Gomez GB, Eakle R, Mukandavire C, Subedar H, et al. Impact and cost-effectiveness of the national scale-up of HIV pre-exposure prophylaxis among female sex workers in South Africa: a modelling analysis. J Int AIDS Soc. 2023;26(2):.

24. Reidy M, Gardiner E, Pretorius C, Glaubius R, Torjesen K, Kripke K. Evaluating the potential impact and cost-effectiveness of dapivirine vaginal ring pre-exposure prophylaxis for HIV prevention. PLoS One. 2019;14(6):e0218710.

25. Glaubius R, Ding Y, Penrose KJ, Hood G, Engquist E, Mellors JW, et al. Dapivirine vaginal ring for HIV prevention: modelling health outcomes, drug resistance and cost-effectiveness. J Int AIDS Soc. 2019;22(5):.

26. Glaubius RL, Hood G, Penrose KJ, Parikh UM, Mellors JW, Bendavid E, et al. Cost-effectiveness of Injectable Preexposure Prophylaxis for HIV Prevention in South Africa. Clin Infect Dis. 2016;63(4):539– 47.

27. Jamieson L, Johnson LF, Nichols BE, Delany-Moretlwe S, Hosseinipour MC, Russell C, et al. Relative cost-effectiveness of long-acting injectable cabotegravir versus oral pre-exposure prophylaxis in South Africa based on the HPTN 083 and HPTN 084 trials: a modelled economic evaluation and threshold analysis. The Lancet HIV.2022; 9: e857–67.

28. Long LC, Girdwood S, Govender K, Meyer-Rath G, Miot J. Cost and outcomes of routine HIV care and treatment: public and private service delivery models covering low-income earners in South Africa. BMC Health Serv Res. 2023;23(1):240.

29. McBain R, Jordan M, Kapologwe N, Kagaayi J, Kiracho E, Nandakumar A. Costing of HIV services, Uganda and United Republic of Tanzania. Bulletin of the World Health Organization 2023; 101 (10): 626– 36.

30. Meyer-Rath G, Rensburg Cv, Larson B, Jamieson L, Rosen S. Revealed willingness-to-pay versus standard cost-effectiveness thresholds: Evidence from the South African HIV Investment Case. PLoS One. 2017;12(10):e0186496.

31. Palanee-Phillips T, Rees HV, Heller KB, Ahmed K, Batting J, Beesham I, et al. High HIV incidence among young women in South Africa: Data from a large prospective study. PLoS One. 2022;17(6):e0269317.

32. Joshi K, Lessler J, Olawore O, Loevinsohn G, Bushey S, Tobian AAR, et al. Declining HIV incidence in sub-Saharan Africa: a systematic review and meta-analysis of empiric data. Journal of the International AIDS Society. 2021; 24: e25818.

33. Risher KA, Cori A, Reniers G, Marston M, Calvert C, Crampin A, et al. Age patterns of HIV incidence in eastern and southern Africa: a modelling analysis of observational population-based cohort studies. Vol. 8, The Lancet HIV 2021; 8: e429–39.

34. Birdthistle I, Kwaro D, Shahmanesh M, Baisley K, Khagayi S, Chimbindi N, et al. Evaluating the impact of DREAMS on HIV incidence among adolescent girls and young women: A population-based cohort study in Kenya and South Africa. PLOS Medicine 2001: e1003837.

35. Sullivan PS, Phaswana-Mafuya N, Baral SD, Valencia R, Zahn R, Dominguez K, et al. HIV prevalence and incidence in a cohort of South African men and transgender women who have sex with men: the Sibanye Methods for Prevention Packages Programme (MP3) project. Journal of the International AIDS Society 2020; 23 (56): e25591.

36. Eaton JW, Brown T, Puckett R, Glaubius R, Mutai K, Bao L, et al. The Estimation and Projection Package Age-Sex Model and the r-hybrid model. AIDS. 2019;33(Supplement 3):S235–44.

37. UNAIDS. UNAIDS Data 2022. Geneva.

38. Kagaayi J, Gray RH, Whalen C, Fu P, Neuhauser D, McGrath JW, et al. Indices to Measure Risk of HIV Acquisition in Rakai, Uganda. Vol. 9, PLoS ONE. 2014; 9(4): e92015.

39. Balkus JE, Brown E, Palanee T, Nair G, Gafoor Z, Zhang J, et al. An Empiric HIV Risk Scoring Tool to Predict HIV-1 Acquisition in African Women. Vol. 72, JAIDS Journal of Acquired Immune Deficiency Syndromes. 2016; 72 (3): 333–43.

40. Moyo RC, Govindasamy D, Manda S, Nyasulu PS. A Prediction Risk Score for HIV among Adolescent Girls and Young Women in South Africa: Identifying those in Need of HIV Pre-Exposure Prophylaxis. A Prediction Risk Score for HIV among Adolescent Girls and Young Women in South Africa: Identifying those in Need of HIV Pre-Exposure Prophylaxis. 0.

